# Protection of Omicron bivalent vaccine, previous infection, and their induced neutralizing antibodies against symptomatic infection with Omicron XBB.1.16 and EG.5.1

**DOI:** 10.1101/2024.05.30.24308254

**Authors:** Shohei Yamamoto, Kouki Matsuda, Kenji Maeda, Tetsuya Mizoue, Kumi Horii, Kaori Okudera, Tomofumi Tan, Yusuke Oshiro, Natsumi Inamura, Takashi Nemoto, Junko S. Takeuchi, Maki Konishi, Haruhito Sugiyama, Nobuyoshi Aoyanagi, Wataru Sugiura, Norio Ohmagari

**Affiliations:** Department of Epidemiology and Prevention, Center for Clinical Sciences, National Center for Global Health and Medicine, Tokyo, Japan; Division of Antiviral Therapy, Joint Research Center for Human Retrovirus Infection, Kagoshima University, Kagoshima, Japan; Department of Refractory Viral Infection, Research Institute, National Center for Global Health and Medicine, Tokyo, Japan; Infection Control Office, Center Hospital of the National Center for the Global Health and Medicine, Tokyo, Japan; Infection Control Office, Kohnodai Hospital of the National Center for the Global Health and Medicine, Chiba, Japan; Department of Laboratory Testing, Center Hospital of the National Center for the Global Health and Medicine, Tokyo, Japan; Department of Academic-Industrial Partnerships Promotion, Center for Clinical Sciences, National Center for Global Health and Medicine, Tokyo, Japan; Center Hospital of the National Center for the Global Health and Medicine, Tokyo, Japan; Kohnodai Hospital of the National Center for the Global Health and Medicine, Chiba, Japan; Center for Clinical Sciences, National Center for Global Health and Medicine, Tokyo, Japan; Disease Control and Prevention Center, National Center for Global Health and Medicine, Tokyo, Japan

**Keywords:** COVID-19, Omicron, bivalent vaccine, previous infection, neutralizing antibody, protection

## Abstract

**Background:** Data are limited on the protective role of the Omicron BA bivalent vaccine, previous infection, and their induced neutralizing antibodies against Omicron XBB.1.16 and EG.5.1 infection.

**Methods:** We conducted a nested case-control analysis among tertiary hospital staff in Tokyo who had received three or more doses of COVID-19 vaccines and donated blood samples in June 2023 (1 month before Omicron XBB.1.16 and EG.5.1 wave). We identified 206 symptomatic cases between June and September 2023 and selected their controls with 1:1 propensity-score matching. We examined the association of vaccination, previous infection, and preinfection live-virus neutralizing antibody titers against Omicron XBB.1.16 and EG.5.1 with the risk of COVID-19 infection.

**Results:** Previous infection during Omicron BA- or XBB-dominant phases was associated with a significantly lower infection risk during the XBB.1.16 and EG.5.1 dominant phase than infection-naïve with 70% and 100% protection, respectively, whereas Omicron BA bivalent vaccination showed no association. Preinfection-neutralizing titers against XBB.1.16 and EG.5.1 were 39% (95%CI: 8–60) and 28% (95%CI: 8–44), respectively, lower in cases than in matched controls. Neutralizing activity against XBB.1.16 and EG.5.1. were somewhat detectable in the sera of individuals with previous infection but barely detectable in those who were infection-naïve and received the Omicron bivalent vaccine.

**Conclusions:** In the era when the Omicron XBB vaccine was unavailable, the Omicron BA bivalent vaccine did not confer the neutralizing activity and protection against Omicron XBB.1.16 and EG.5.1 symptomatic infection. The previous infection afforded neutralizing titers and protection against symptomatic infection with these variants.

**Main points:** In the era when the Omicron XBB vaccine was unavailable, the Omicron BA bivalent vaccine did not confer the neutralizing activity and protection against Omicron XBB.1.16 and EG.5.1 symptomatic infection. The previous infection afforded neutralizing titers and protection against symptomatic infection with these variants.

## Introduction

As of 2024, four years after the initial outbreak, the COVID-19 pandemic is still ongoing due to the persistent mutation cycle of the SARS-CoV-2. In late 2020, clinical trials showed that the COVID-19 vaccination was highly effective in lowering the risk of SARS-CoV-2 infection and severe outcomes [1, 2]. In late 2021, COVID-19 cases rapidly increased due to the Omicron BA subvariants among the vaccinated population. In 2022, a subsequent updated Omicron BA bivalent vaccine lowered the risk of Omicron BA infections [3]. In early- to mid-2023, the Omicron XBB subvariants (XBB.1.5, XBB.1.16, and EG.5), with multiple spike protein mutations compared to earlier BA subvariants [4–6], dominated worldwide. Until the Omicron XBB vaccine became available in September 2023, people had to rely on immunity acquired by existing vaccines or prior infections against Omicron XBB subvariants.

In immunological studies, both Omicron bivalent BA vaccines and Omicron BA infection elicited neutralizing activity against XBB.1.5, XBB.1.16, and EG.5 subvariants, albeit to a limited extent [7–9]. In epidemiological studies, while the Omicron BA bivalent vaccine or previous Omicron BA infection were reported to confer moderate protection against the Omicron XBB.1.5 infection [3, 10, 11], the evidence regarding the protection against Omicron XBB.1.16 and EG.5 infection is limited. In a cohort study of 51,017 U.S. healthcare workers, the previous Omicron infection, but not the Omicron bivalent vaccine, was associated with a lower risk of subsequent infection when Omicron XBB.1.16 and EG.5 subvariants were dominant [12]. Quantitative association between vaccine- or infection-acquired neutralizing activity against Omicron XBB.1.16 and EG.5 and the risk of infection with these variants remains elusive.

In June 2023, when a month before the Omicron XBB.1.16 and EG.5.1 epidemic in Japan (July to September 2023), we performed a serological survey among the staff of the National Center for Global Health and Medicine (NCGM), Tokyo, and stored blood samples. This situation prompted us to investigate whether the Omicron bivalent vaccine and previous infection could confer protection against Omicron XBB.1.16 and EG.5.1 infection and its induced neutralizing antibody titers could correlate with infection protection.

Here, we examined the protection of the Omicron bivalent vaccine and previous infection against Omicron XBB.1.16 and EG.5.1 infection and compared the live-virus and preinfection-neutralizing antibody titers between infected cases and controls in the nested case-control study of recipients with three or more doses of COVID-19 historical monovalent or Omicron BA bivalent vaccines.

## Methods

### Study setting

A repeat serological study was conducted at the NCGM in Japan in July 2020 to monitor the spread of SARS-CoV-2 infection among staff during the COVID-19 epidemic. The details of this study have been reported elsewhere [13–15]. In summary, we have completed eight serosurveys as of June 2023, where we measured anti-SARS-CoV-2 nucleocapsid-(all serosurveys) and spike-protein antibodies (from the second serosurvey onward) for all the participants using both Abbott and Roche assays, stored serum samples at −80°C, and collected information on COVID-19–related factors (vaccination, occupational infection risk, infection prevention practices, behavioral factors, etc.) via a questionnaire. The self-reported vaccination status was validated using objective information from the NCGM Labor Office. Written informed consent was obtained from all the participants. This study was approved by the NCGM Ethics Committee (approval number: NCGM-G-003598).

### Case-Control Selection

We conducted a nested case-control study among the staff who participated in the eighth survey conducted in June 2023 and had received three or more doses of the mRNA COVID-19 vaccine (any of the patterns of historical monovalent vaccine, Omicron BA.1 and wild-type bivalent vaccine, and Omicron BA.4/5 and wild-type bivalent vaccine) manufactured by Pfizer or Moderna (**Figure S1**). Of the 2,569 participants, 2,409 received three or more doses of the mRNA COVID-19 vaccines and donated blood samples. Of those, we excluded 16 participants who lacked information on covariates: body mass index (n=10), alcohol drinking status (n=2), living arrangement status (n=5), adherence to infection prevention practice (n=3), and infection risk behaviors (n=2). We further excluded 19 participants with insufficient volume of serum sample (<100μL), leaving 2374 participants as the base population.

We followed the participants for COVID-19 incidence using the COVID-19 patient records documented by the NCGM Hospital Infection Prevention and Control Unit. As per the NCGM rule, staff should undergo PCR or antigen test for COVID-19 when they have COVID-19-compatible symptoms, and if it tests positive, they must report the results to the NCGM Hospital Infection Prevention and Control Unit. During the follow-up (June to September 2023), we identified 217 COVID-19 patients. We defined cases as symptomatic SARS-CoV-2 infection. Participants infected after additional vaccination during follow-up were considered cases if the infection occurred within 14 days after the vaccination, assuming they were not sufficiently immunized with the additional booster until then. After excluding 11 asymptomatic patients, leaving 206 were included as cases (**Figure S1**). We selected a control for each case using propensity score matching to compare preinfection anti-spike antibody titers. The details of the case-control matching algorithm are described in **supplemental text 1**. We randomly selected 50 pairs of these matched pairs and measured live virus–neutralizing antibody titers to compare neutralizing antibodies between the groups.

### Antibody Testing

We measured neutralizing activity against Wild-type, Omicron XBB.1.16, and Omicron EG.5.1 in the sera of patients and controls by quantifying the serum-mediated suppression of the cytopathic effect of each SARS-CoV-2 strain in HeLa_hACE2-TMPRSS2_ cells [16, 17]. The details of the measurement methods are described in supplemental text 2.

We assessed anti-SARS-CoV-2 antibodies in all the participants at baseline and retrieved data for the case-control pairs. We quantitatively measured the levels of antibodies against the receptor-binding domain (RBD) of the SARS-CoV-2 spike protein using the AdviseDx SARS-CoV-2 IgG II assay (Abbott) (i.e., anti-RBD immunoglobulin [Ig] G) and Elecsys^®^ Anti-SARS-CoV-2 S (Roche) (i.e., anti-RBD total). We also qualitatively measured antibodies against the SARS-CoV-2 nucleocapsid (N) protein using the SARS-CoV-2 IgG assay (Abbott) and Elecsys^®^ Anti-SARS-CoV-2 (Roche).

### Previous infection status at baseline

Previous infection was defined as a self-reported history of COVID-19 (confirmed against in-house COVID-19 registry) at baseline or anti-N seropositive with any of the two assays (Roche ≥1.0 COI or Abbott ≥ 1.40 S/C) at any of the first (July 2020) through eighth (June 2023: baseline) surveys. We defined participants with no history of COVID-19 but seropositive on N antibodies as undiagnosed infection [18]. We defined phases of previous infection referring to the timing of the last diagnosis: Pre-Omicron (February 2020 to December 2021), Omicron BA (January 2022 to March 2023), and Omicron XBB (April 2024 to June 2024).

### Statistical analysis

We used conditional logistic regression while accounting for the matched design to examine the association of vaccination status (doses of any COVID-19 vaccines and dose of Omicron BA vaccines) and previous infection status with COVID-19 infection risk. We used a generalized estimating equation (GEE) with group assignment (case or control) and a robust variance estimator to compare the interval from the last vaccination or COVID-19 diagnosis to baseline blood sampling. To examine the difference in preinfection antibody levels between cases and controls, we compared the log-transformed titers of neutralizing (Wild-type, Omicron XBB.1.16 and Omicron EG.5.1) and anti-RBD (IgG and total) antibodies between matched pairs using a GEE model with group assignment and a robust variance estimator. Then, we back-transformed and presented these values as geometric mean titers (GMTs) with 95% confidence intervals (CIs). We repeated the GEE analysis by restricting matched pairs to infection-naïve pairs at baseline (i.e., both case and matched controls had no history of COVID-19 and were negative on anti-N assays) as sensitivity analysis. We used the Kruskal–Wallis test to compare the neutralizing titers across vaccination status (historical monovalent vaccine only or the historical monovalent plus Omicron bivalent vaccines) and previous infection status (infection-naïve or previously infected). To examine the difference in neutralizing titers across the timing of previous infection, we used a linear regression model while adjusting age, sex, a history of Omicron bivalent vaccination, and the interval between the last vaccination and blood sampling. For the analyses of neutralizing antibody titers, values below the limit of detection (LOD) (NT_50_ < 40) were given the LOD value. Statistical analyses were performed using Stata version 18.0 (StataCorp LLC), and graphics were generated using GraphPad Prism 9 (GraphPad, Inc.). All P-values were 2-sided, and the statistical significance was set at P<0.05.

## Results

### Distribution of circulating SARS-CoV-2 variants during follow-up

**Figure 1** shows the distribution of SARS-CoV-2 lineages in Japan during the study period (June to September 2023), analyzed using all domestic genome sequences registered in the GISAID EpiCov database (https://gisaid.org). During the study period, 27,899 samples were extracted for sequences, and the most frequent subvariants were Omicron XBB variants with a relative frequency of 91%. According to subvariants, the most frequent subvariants were Omicron XBB.1.16 (23%), following Omicron EG.5 (22%). From June to September 2023, the relative frequency of Omicron XBB.1.16 decreased (28% to 18%), while that of Omicron EG.5 increased (13% to 31%).

**Figure 1.**
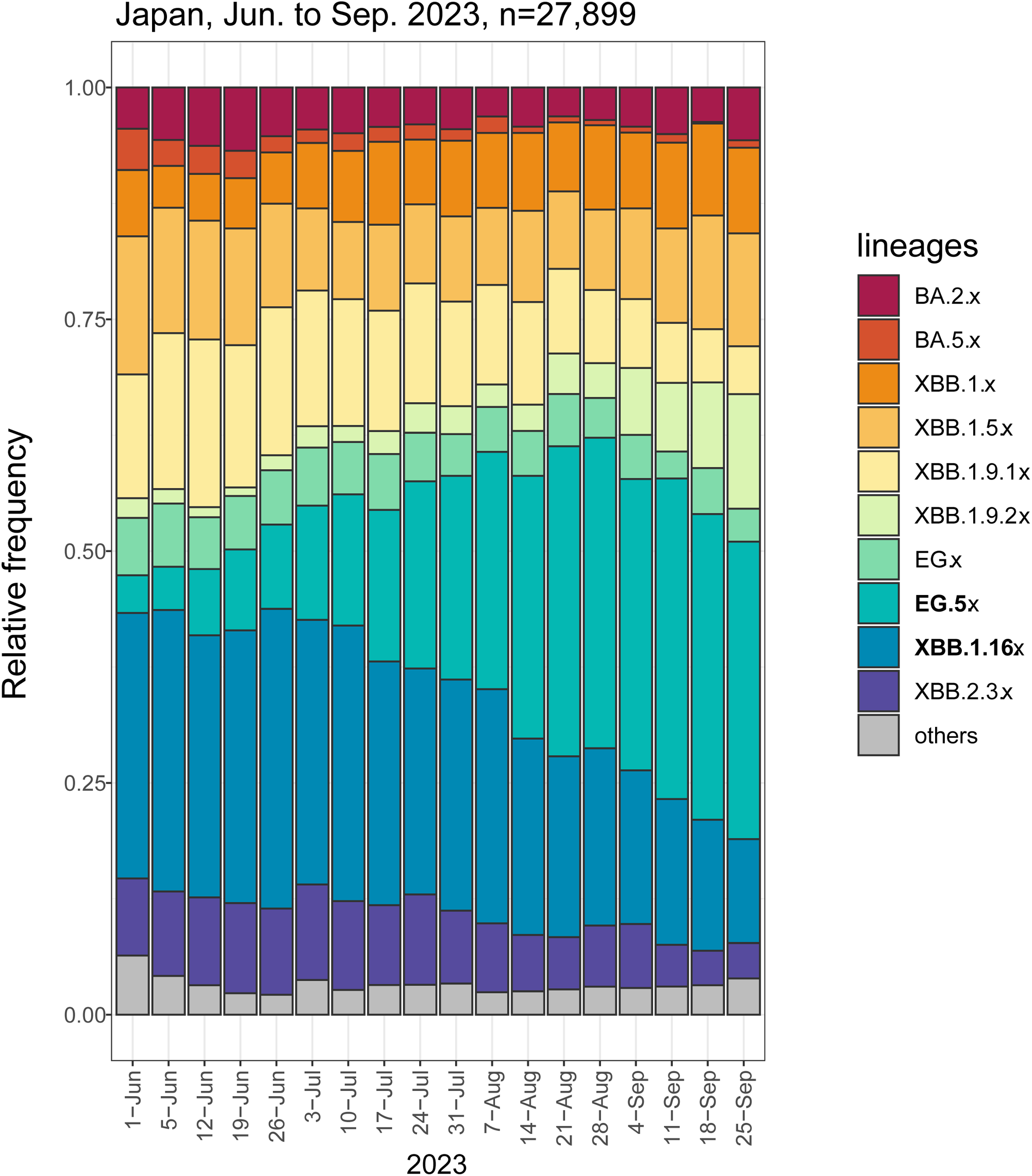
Relative frequency of circulating SARS-CoV-2 variants from June to September 2023 in Japan. The distribution of SARS-CoV-2 lineages in Japan during the study period (June – September 2023) was analyzed using all domestic genome sequences registered in the GISAID EpiCov database (https://gisaid.org). For lineage analysis, the extracted sequences (n=27,899) were applied to the Phylogenetic Assignment of Named Global Outbreak Lineages (PANGOLIN) version 4.3.1 with pangolin-data 1.25.1.

### Baseline characteristics before and after propensity score matching

We ascertained 206 symptomatic breakthrough infection cases during the follow-up in the before-matching cohort, with an incidence rate of 12.7 per 10000 person-days. Cases were younger and more likely to be female and nurses than the controls in the before-matching cohort (**Table 1**). After propensity matching with a 1:1 ratio, the 206 matched pairs were well-balanced regarding all the baseline characteristics.

**Table 1.**
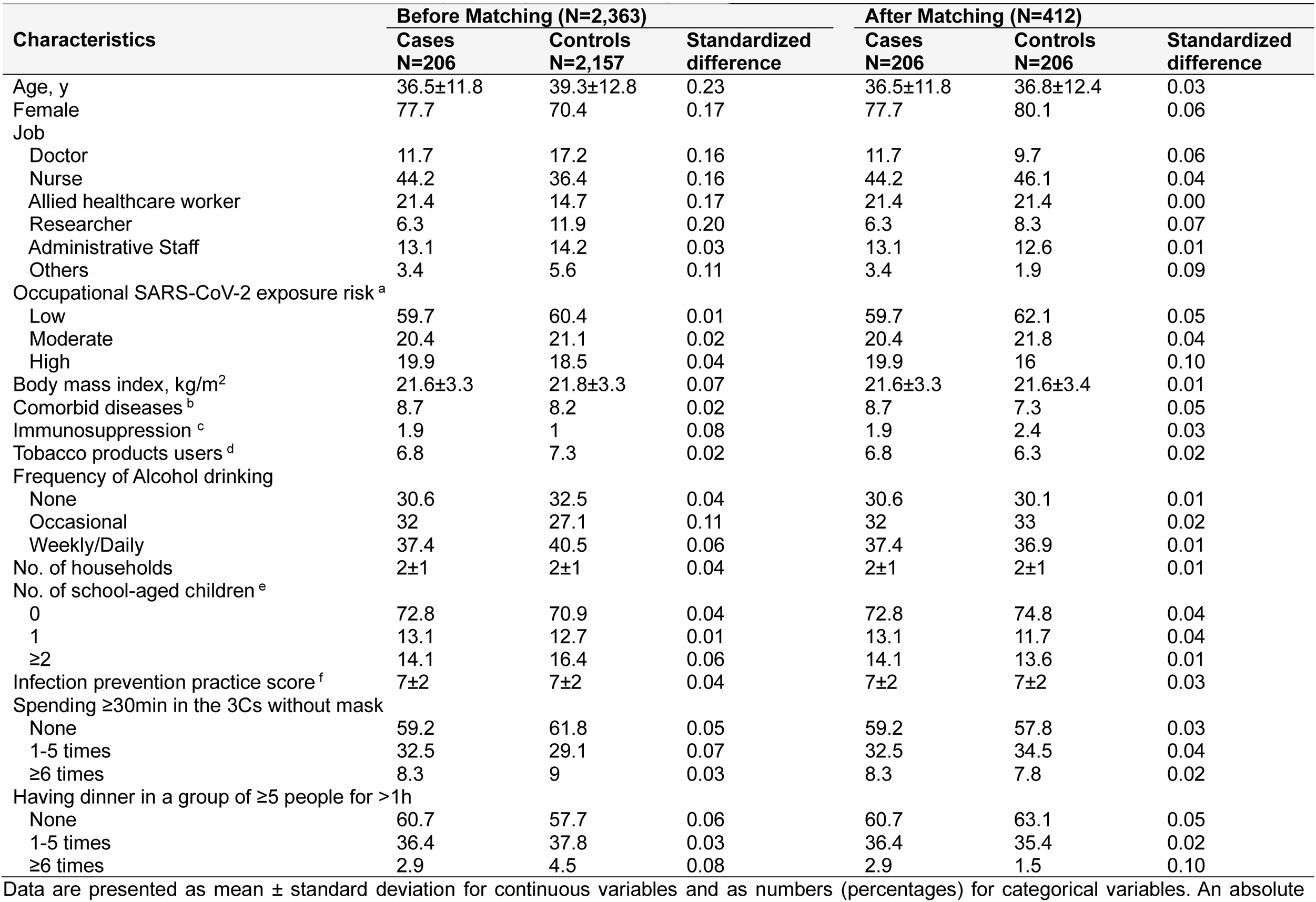
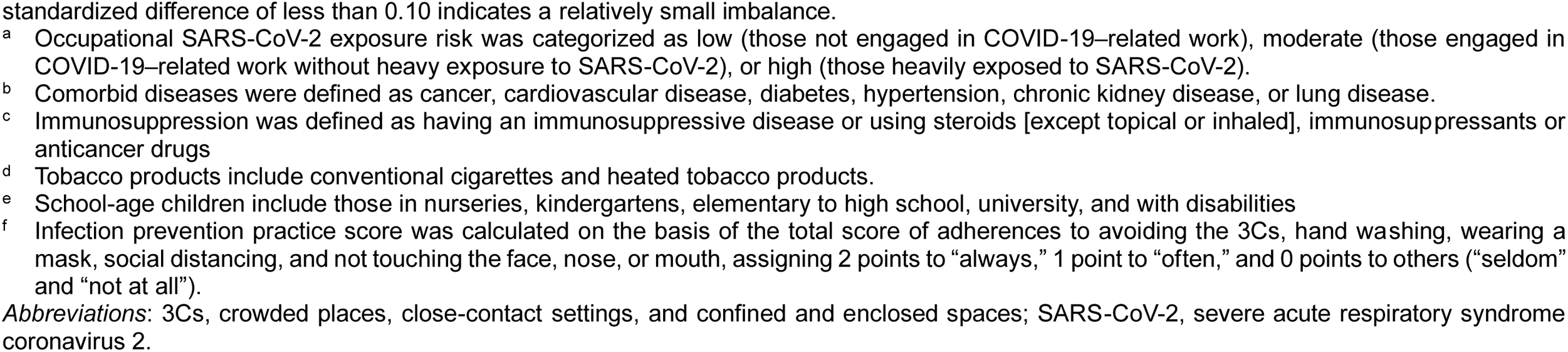
Baseline characteristics before and after propensity score matching.

| Characteristics | Before Matching (N=2,363) |  |  | After Matching (N=412) |  |  |
| --- | --- | --- | --- | --- | --- | --- |
|  | Cases<br>N=206 | Controls<br>N=2,157 | Standardized<br>difference | Cases<br>N=206 | Controls<br>N=206 | Standardized<br>difference |
| Age, y | 36.5±11.8 | 39.3±12.8 | 0.23 | 36.5±11.8 | 36.8±12.4 | 0.03 |
| Female | 77.7 | 70.4 | 0.17 | 77.7 | 80.1 | 0.06 |
| Job |  |  |  |  |  |  |
| Doctor | 11.7 | 17.2 | 0.16 | 11.7 | 9.7 | 0.06 |
| Nurse | 44.2 | 36.4 | 0.16 | 44.2 | 46.1 | 0.04 |
| Allied healthcare worker | 21.4 | 14.7 | 0.17 | 21.4 | 21.4 | 0.00 |
| Researcher | 6.3 | 11.9 | 0.20 | 6.3 | 8.3 | 0.07 |
| Administrative Staff | 13.1 | 14.2 | 0.03 | 13.1 | 12.6 | 0.01 |
| Others | 3.4 | 5.6 | 0.11 | 3.4 | 1.9 | 0.09 |
| Occupational SARS-CoV-2 exposure risk <sup>a</sup> |  |  |  |  |  |  |
| Low | 59.7 | 60.4 | 0.01 | 59.7 | 62.1 | 0.05 |
| Moderate | 20.4 | 21.1 | 0.02 | 20.4 | 21.8 | 0.04 |
| High | 19.9 | 18.5 | 0.04 | 19.9 | 16 | 0.10 |
| Body mass index, kg/m<sup>2</sup> | 21.6±3.3 | 21.8±3.3 | 0.07 | 21.6±3.3 | 21.6±3.4 | 0.01 |
| Comorbid diseases <sup>b</sup> | 8.7 | 8.2 | 0.02 | 8.7 | 7.3 | 0.05 |
| Immunosuppression <sup>c</sup> | 1.9 | 1 | 0.08 | 1.9 | 2.4 | 0.03 |
| Tobacco products users <sup>d</sup> | 6.8 | 7.3 | 0.02 | 6.8 | 6.3 | 0.02 |
| Frequency of Alcohol drinking |  |  |  |  |  |  |
| None | 30.6 | 32.5 | 0.04 | 30.6 | 30.1 | 0.01 |
| Occasional | 32 | 27.1 | 0.11 | 32 | 33 | 0.02 |
| Weekly/Daily | 37.4 | 40.5 | 0.06 | 37.4 | 36.9 | 0.01 |
| No. of households | 2±1 | 2±1 | 0.04 | 2±1 | 2±1 | 0.01 |
| No. of school-aged children <sup>e</sup> |  |  |  |  |  |  |
| 0 | 72.8 | 70.9 | 0.04 | 72.8 | 74.8 | 0.04 |
| 1 | 13.1 | 12.7 | 0.01 | 13.1 | 11.7 | 0.04 |
| ≥2 | 14.1 | 16.4 | 0.06 | 14.1 | 13.6 | 0.01 |
| Infection prevention practice score <sup>f</sup> | 7±2 | 7±2 | 0.04 | 7±2 | 7±2 | 0.03 |
| Spending ≥30min in the 3Cs without mask |  |  |  |  |  |  |
| None | 59.2 | 61.8 | 0.05 | 59.2 | 57.8 | 0.03 |
| 1-5 times | 32.5 | 29.1 | 0.07 | 32.5 | 34.5 | 0.04 |
| ≥6 times | 8.3 | 9 | 0.03 | 8.3 | 7.8 | 0.02 |
| Having dinner in a group of ≥5 people for >1h |  |  |  |  |  |  |
| None | 60.7 | 57.7 | 0.06 | 60.7 | 63.1 | 0.05 |
| 1-5 times | 36.4 | 37.8 | 0.03 | 36.4 | 35.4 | 0.02 |
| ≥6 times | 2.9 | 4.5 | 0.08 | 2.9 | 1.5 | 0.10 |

Data are presented as mean ± standard deviation for continuous variables and as numbers (percentages) for categorical variables. An absolute
standardized difference of less than 0.10 indicates a relatively small imbalance.
- <sup>a</sup> Occupational SARS-CoV-2 exposure risk was categorized as low (those not engaged in COVID-19–related work), moderate (those engaged in COVID-19–related work without heavy exposure to SARS-CoV-2), or high (those heavily exposed to SARS-CoV-2). - <sup>b</sup> Comorbid diseases were defined as cancer, cardiovascular disease, diabetes, hypertension, chronic kidney disease, or lung disease. - <sup>c</sup> Immunosuppression was defined as having an immunosuppressive disease or using steroids [except topical or inhaled], immunosuppressants or anticancer drugs - <sup>d</sup> Tobacco products include conventional cigarettes and heated tobacco products. - <sup>e</sup> School-age children include those in nurseries, kindergartens, elementary to high school, university, and with disabilities - <sup>f</sup> Infection prevention practice score was calculated on the basis of the total score of adherences to avoiding the 3Cs, hand washing, wearing a mask, social distancing, and not touching the face, nose, or mouth, assigning 2 points to “always,” 1 point to “often,” and 0 points to others (“seldom” and “not at all”).
*Abbreviations:* 3Cs, crowded places, close-contact settings, and confined and enclosed spaces; SARS-CoV-2, severe acute respiratory syndrome coronavirus 2.

### Vaccination and previous infection statuses and risk of COVID-19 infection

The number of existing mRNA vaccinations and those of the Omicron BA bivalent vaccination were not associated with the risk of COVID-19 infection (**Table 2**). In the analysis of any mRNA vaccines, the OR (95% CI) of 3 to 6 doses against infection were 1 (reference), 1.01 (0.62 to 1.63), 0.78 (0.47 to 1.30), and 1.04 (0.32 to 3.39), respectively. For the analysis of Omicron BA bivalent vaccines, the OR (95% CI) of no vaccination (monovalent vaccine only), 1-dose, and 2-dose were 1 (reference), 0.79 (0.52 to 1.20), 1.40 (0.32 to 6.06), respectively. The mean interval days between the last vaccination and baseline blood sampling were not statistically different between cases and controls (287 v.s. 274 days).

**Table 2.**
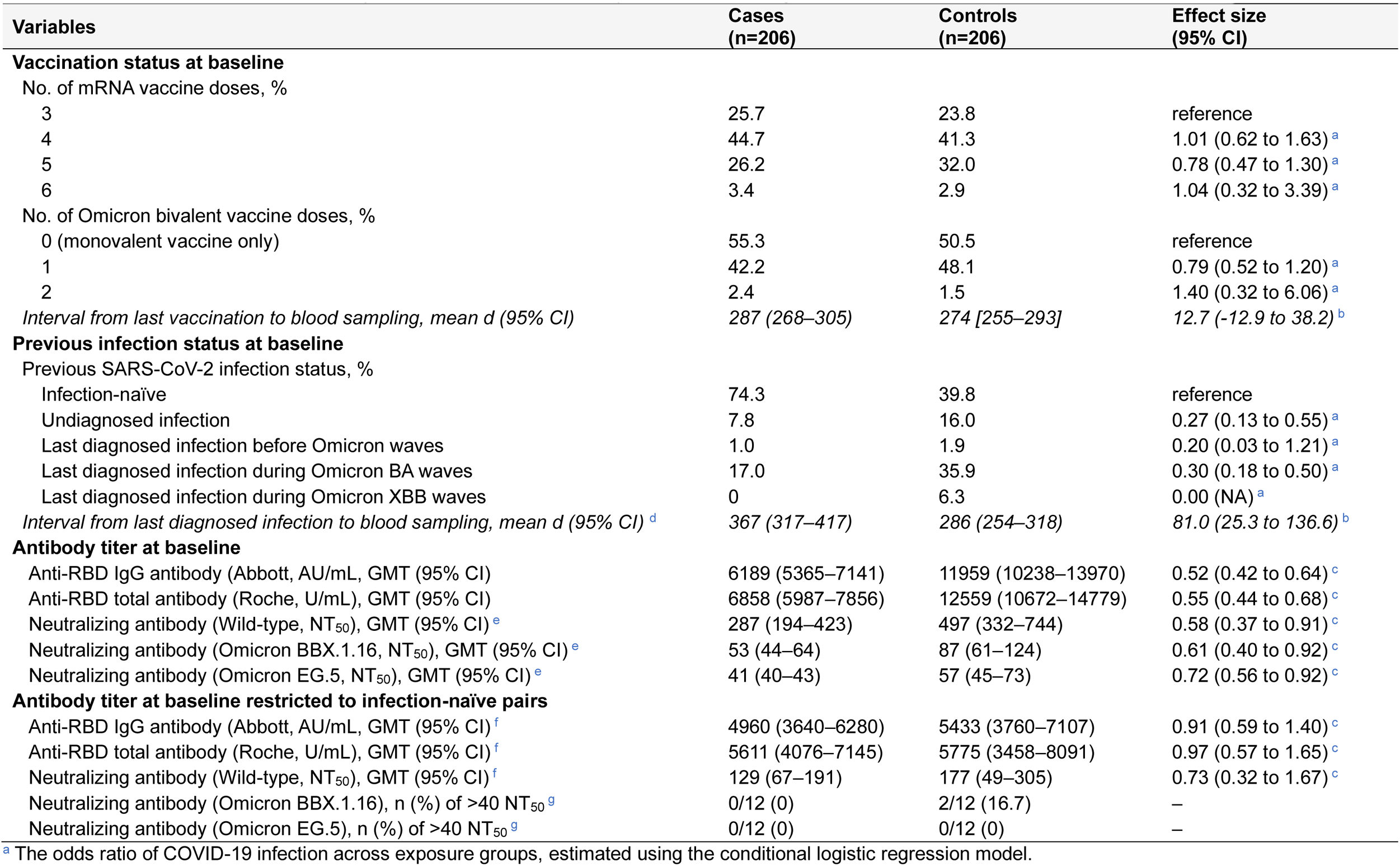
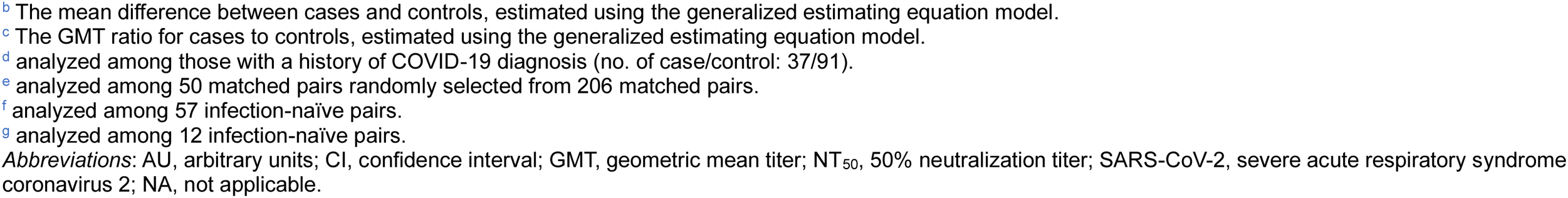
Association of vaccination status, previous infection status, and preinfection antibody titers with the risk of COVID-19 infection.

| Variables | Cases<br>(n=206) | Controls<br>(n=206) | Effect size<br>(95% CI) |
| --- | --- | --- | --- |
| **Vaccination status at baseline** |  |  |  |
| No. of mRNA vaccine doses, % |  |  |  |
| 3 | 25.7 | 23.8 | reference |
| 4 | 44.7 | 41.3 | 1.01 (0.62 to 1.63) <sup>a</sup> |
| 5 | 26.2 | 32.0 | 0.78 (0.47 to 1.30) <sup>a</sup> |
| 6 | 3.4 | 2.9 | 1.04 (0.32 to 3.39) <sup>a</sup> |
| No. of Omicron bivalent vaccine doses, % |  |  |  |
| 0 (monovalent vaccine only) | 55.3 | 50.5 | reference |
| 1 | 42.2 | 48.1 | 0.79 (0.52 to 1.20) <sup>a</sup> |
| 2 | 2.4 | 1.5 | 1.40 (0.32 to 6.06) <sup>a</sup> |
| *Interval from last vaccination to blood sampling, mean d (95% CI)* | *287 (268–305)* | *274 [255–293]* | *12.7 (-12.9 to 38.2) <sup>b</sup>* |
| **Previous infection status at baseline** |  |  |  |
| Previous SARS-CoV-2 infection status, % |  |  |  |
| Infection-naïve | 74.3 | 39.8 | reference |
| Undiagnosed infection | 7.8 | 16.0 | 0.27 (0.13 to 0.55) <sup>a</sup> |
| Last diagnosed infection before Omicron waves | 1.0 | 1.9 | 0.20 (0.03 to 1.21) <sup>a</sup> |
| Last diagnosed infection during Omicron BA waves | 17.0 | 35.9 | 0.30 (0.18 to 0.50) <sup>a</sup> |
| Last diagnosed infection during Omicron XBB waves | 0 | 6.3 | 0.00 (NA) <sup>a</sup> |
| *Interval from last diagnosed infection to blood sampling, mean d (95% CI) <sup>d</sup>* | *367 (317–417)* | *286 (254–318)* | *81.0 (25.3 to 136.6) <sup>b</sup>* |
| **Antibody titer at baseline** |  |  |  |
| Anti-RBD IgG antibody (Abbott, AU/mL, GMT (95% CI) | 6189 (5365–7141) | 11959 (10238–13970) | 0.52 (0.42 to 0.64) <sup>c</sup> |
| Anti-RBD total antibody (Roche, U/mL), GMT (95% CI) | 6858 (5987–7856) | 12559 (10672–14779) | 0.55 (0.44 to 0.68) <sup>c</sup> |
| Neutralizing antibody (Wild-type, NT<sub>50</sub>), GMT (95% CI) <sup>e</sup> | 287 (194–423) | 497 (332–744) | 0.58 (0.37 to 0.91) <sup>c</sup> |
| Neutralizing antibody (Omicron BBX.1.16, NT<sub>50</sub>), GMT (95% CI) <sup>e</sup> | 53 (44–64) | 87 (61–124) | 0.61 (0.40 to 0.92) <sup>c</sup> |
| Neutralizing antibody (Omicron EG.5, NT<sub>50</sub>), GMT (95% CI) <sup>e</sup> | 41 (40–43) | 57 (45–73) | 0.72 (0.56 to 0.92) <sup>c</sup> |
| **Antibody titer at baseline restricted to infection-naïve pairs** |  |  |  |
| Anti-RBD IgG antibody (Abbott, AU/mL, GMT (95% CI) <sup>f</sup> | 4960 (3640–6280) | 5433 (3760–7107) | 0.91 (0.59 to 1.40) <sup>c</sup> |
| Anti-RBD total antibody (Roche, U/mL), GMT (95% CI) <sup>f</sup> | 5611 (4076–7145) | 5775 (3458–8091) | 0.97 (0.57 to 1.65) <sup>c</sup> |
| Neutralizing antibody (Wild-type, NT<sub>50</sub>), GMT (95% CI) <sup>f</sup> | 129 (67–191) | 177 (49–305) | 0.73 (0.32 to 1.67) <sup>c</sup> |
| Neutralizing antibody (Omicron BBX.1.16), n (%) of >40 NT<sub>50</sub> <sup>g</sup> | 0/12 (0) | 2/12 (16.7) | – |
| Neutralizing antibody (Omicron EG.5), n (%) of >40 NT<sub>50</sub> <sup>g</sup> | 0/12 (0) | 0/12 (0) | – |

<sup>a</sup> The odds ratio of COVID-19 infection across exposure groups, estimated using the conditional logistic regression model.
- <sup>b</sup> The mean difference between cases and controls, estimated using the generalized estimating equation model. - <sup>c</sup> The GMT ratio for cases to controls, estimated using the generalized estimating equation model. - <sup>d</sup> analyzed among those with a history of COVID-19 diagnosis (no. of case/control: 37/91). - <sup>e</sup> analyzed among 50 matched pairs randomly selected from 206 matched pairs. - <sup>f</sup> analyzed among 57 infection-naïve pairs. - <sup>g</sup> analyzed among 12 infection-naïve pairs.
*Abbreviations:* AU, arbitrary units; CI, confidence interval; GMT, geometric mean titer; NT<sub>50</sub>, 50% neutralization titer; SARS-CoV-2, severe acute respiratory syndrome coronavirus 2; NA, not applicable.

Previous infection at the Omicron BA or XBB phase, but not at the pre-Omicron phase, was significantly associated with a lower risk of COVID-19 infection during the follow-up (**Table 2**). Compared to infection-naïve, the OR (95% CI) of previous infection at pre-Omicron and Omicron BA waves against infection were 0.20 (0.03 to 1.21) and 0.30 (0.18 to 0.50), respectively. No COVID-19 infection occurred in the group of previous infection at the Omicron XBB wave (i.e., OR=0.00). Undiagnosed infection was also associated with a lower risk of infection than infection-naïve, with an OR (95% CI) of 0.27 (0.13 to 0.55). Among the participants with a history of COVID-19, the interval between the last infection and baseline blood sampling was statistically longer in cases than in controls, with a mean difference (95% CI) of 81 (25 to 137) days.

### Preinfection antibody titers between the cases and matched controls

The GMT of preinfection-neutralizing antibodies against Wild-type, Omicron XBB.1.16, and EG.5.1 were 377, 68, and 49, and their detection rate (>40 NT_50_) were 89%, 28%, and 14%, respectively, among total samples of cases and controls (**Figure S2**).

Preinfection anti-RBD and neutralizing antibody titers were lower in cases than in controls. The GEE-predicted GMTs (95% CI) of the anti-RBD IgG antibody on Abbott assay (AU/ml) was 6189 (5365–7141) for cases and 11959 (10238–13970) for controls with a predicted case-to-control ratio of the titers of 0.52 (95% CI: 0.42– 0.64) (**Table 2** and **Figure 2**). The GMTs (95% CI) of the anti-RBD total antibody on Roche assay (U/mL) were 6858 (5987–7856) for cases and 12559 (10672–14779) with a ratio of 0.55 (95% CI: 0.44–0.68). The predicted neutralizing antibody GMTs (95% CI) against Wild-type (NT_50_) were 287 (194–423) for cases and 497 (332–744) for controls, with a ratio of 0.58 (95% CI: 0.37–0.91). The detection rate of neutralization (i.e., ≥40 NT_50_) against Omicron XBB.1.16 was lower in cases than in controls (20% vs. 36%), and the GMTs (95% CI) of Omicron XBB.1.16 were 53 (44–64) for cases and 87 (61–124) for controls. The rate of neutralization detection against Omicron EG.5.1 was lower in cases than in controls (6% vs. 22%), and the GMTs (95% CI) of Omicron EG.5.1 were 41 (40–43) for cases and 57 (45–73) for controls.

**Figure 2.**
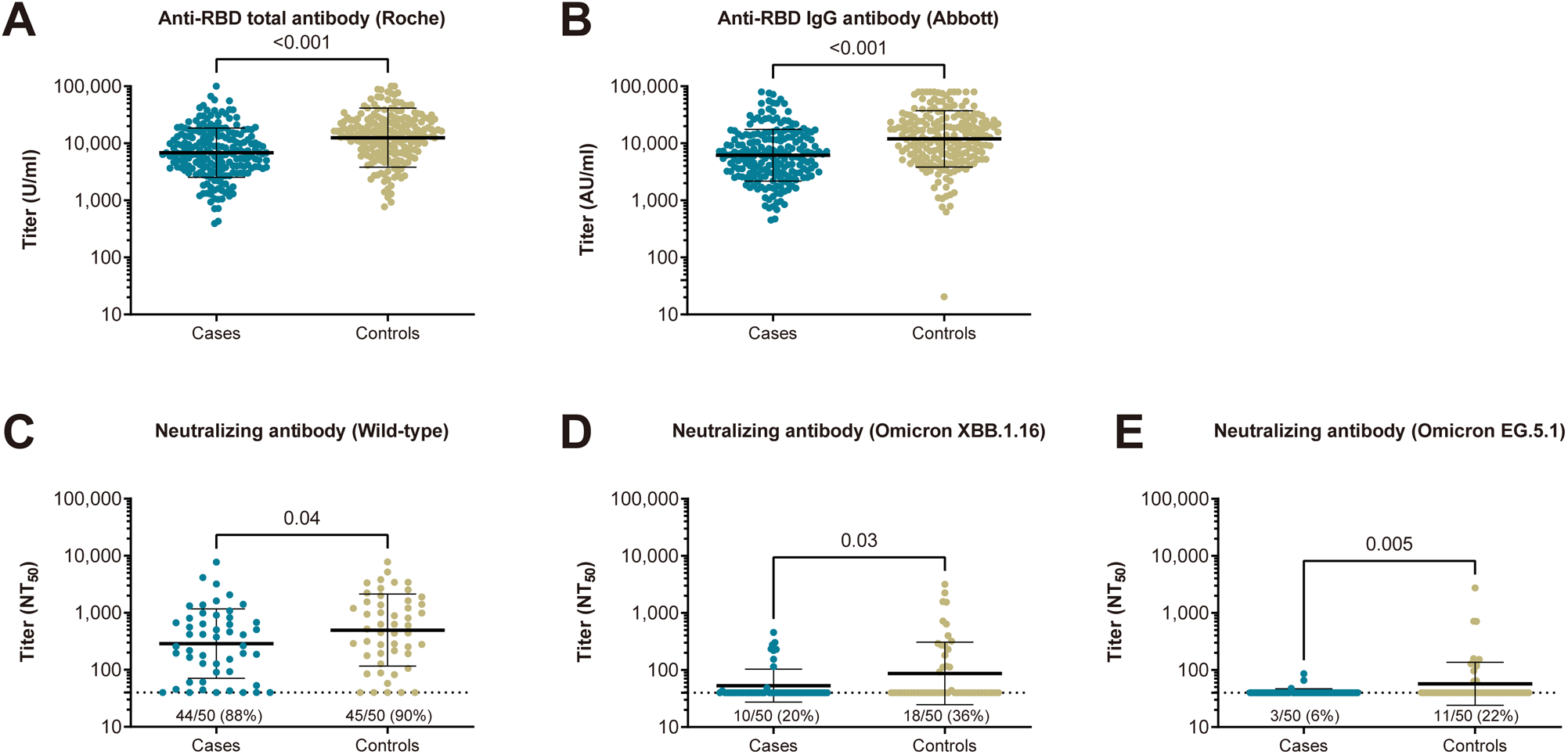
Comparison of the preinfection live-virus neutralizing and anti-RBD antibody titers between propensity-score matched cases and controls. Preinfection anti-RBD IgG antibody titers were measured using the Abbott reagent (**A**), and anti-RBD total antibody titers were measured with Roche reagent (**B**) among 206 cases with breakthrough infection and 206 matched controls. In addition, the live-virus neutralizing antibody titers against Wild-type (**C**), Omicron XBB.1.16 (**D**), and Omicron EG.5.1 (**E**) among the 50 matched pairs were randomly selected from 206 matched pairs. In each panel, the horizontal bars indicate the geometric mean titers, and the I-shaped bars indicate the geometric standard deviations. The limit of detection (LOD) of the neutralizing assay is 40, as shown by dashed horizontal lines. The number (%) of the upper LOD is denoted above the X-axis. Abbreviations: AU, arbitrary units; IgG, immunoglobulin G; NT_50_, 50% neutralizing titer; RBD, receptor-binding domain

The sensitivity analyses restricting to infection-naïve matched pairs yield that the difference in preinfection anti-RBD and neutralizing antibody titers between cases and controls were attenuated and no longer statistically significant. No infection-naïve cases detected neutralization against Omicron XBB.1.16 and EG.5.1.

### Neutralizing antibody titers across statuses of Omicron bivalent vaccination and previous infection

There were no substantial differences in preinfection-neutralizing antibody titers against Wild-type, Omicron XBB.1.16, and Omicron EG.5.1 between individuals irrespective of their history of Omicron bivalent vaccination. Neutralization against Omicron XBB.1.16 and EG.5.1 was not detected (i.e., <40 NT_50_) in all serums from infection-naïve individuals who never received the Omicron bivalent vaccine. Among infection-naïve individuals with a history of Omicron bivalent vaccine, only 21% and 4% had detectable neutralizing titers against XBB.1.16 and EG.5.1, respectively.

Previously infected individuals had higher preinfection-neutralizing antibody titers against Wild-type, Omicron XBB.1.16, and Omicron EG.5.1 than infection-naïve individuals (**Figure 3**). Irrespective of the previous infection phases, Wild-type neutralizing titers were higher in those with previous infection than those with infection-naïve (**Figure 4**). Those infected during Omicron XBB periods had the highest neutralizing antibody titers against Omicron XBB.1.16 and EG.5.1 compared to those infected in Omicron BA or earlier periods. Those infected in the Omicron BA period had statistically higher neutralizing titers against Omicron XBB.1.16 than those with infection-naïve, whereas not in titers against Omicron EG.5.1.

**Figure 3.**
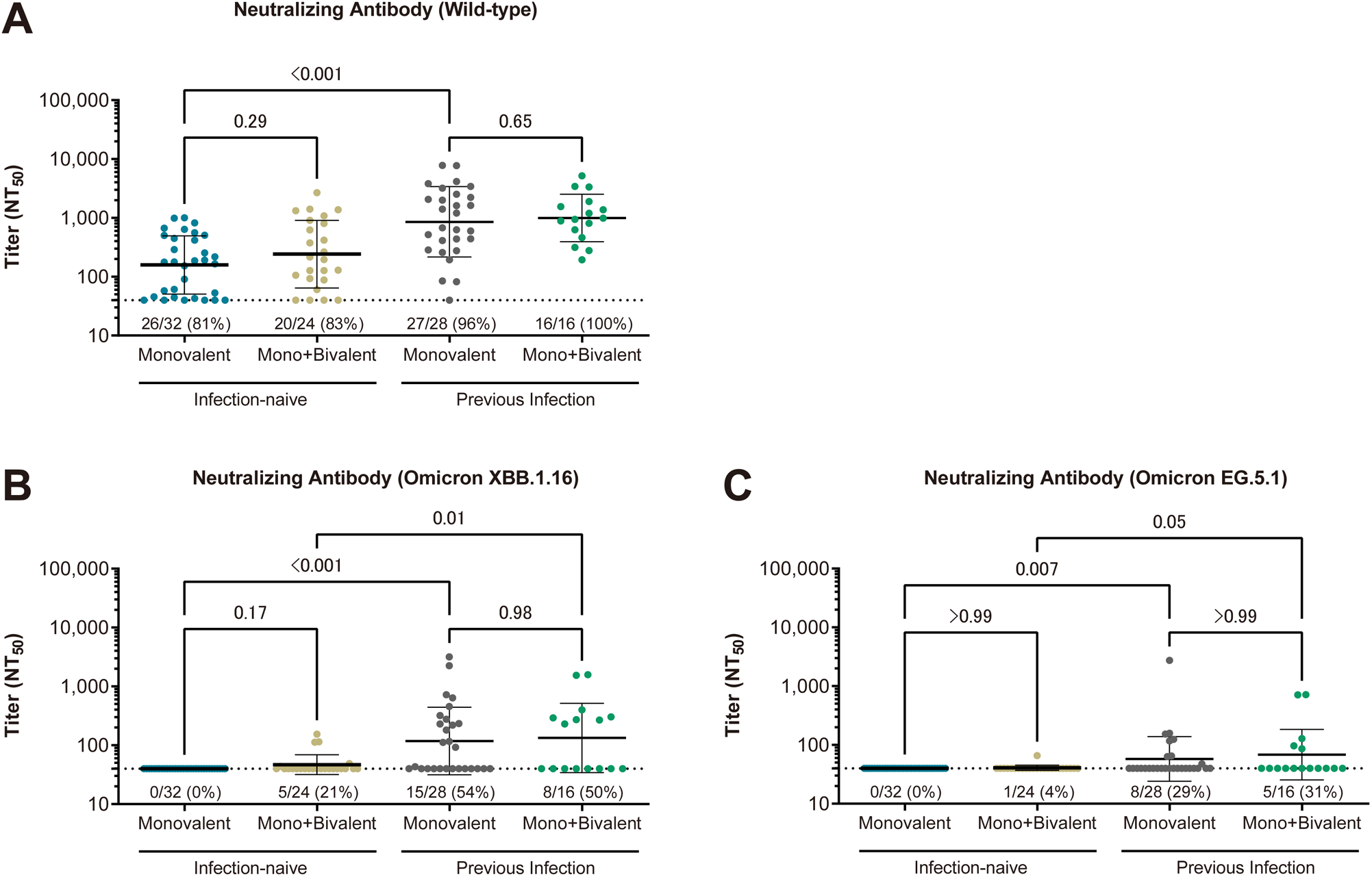
Comparison of the preinfection live-virus neutralizing antibody titers across the histories of Omicron bivalent vaccine and prior SARS-CoV-2 infection. Shown are the live-virus neutralizing antibody titers against Wild-type (**A**), Omicron XBB.1.16 (**B**), and Omicron EG.5.1 (**C**) among the 100 samples from 50 matched pairs. In each panel, the horizontal bars indicate the geometric mean titers, and the I-shaped bars indicate the geometric standard deviations. The limit of detection (LOD) of the neutralizing assay is 40, as shown by dashed horizontal lines. The number (%) of the upper LOD is denoted above the X-axis. Abbreviations: NT_50_, 50% neutralizing titer; RBD, receptor-binding domain

**Figure 4.**
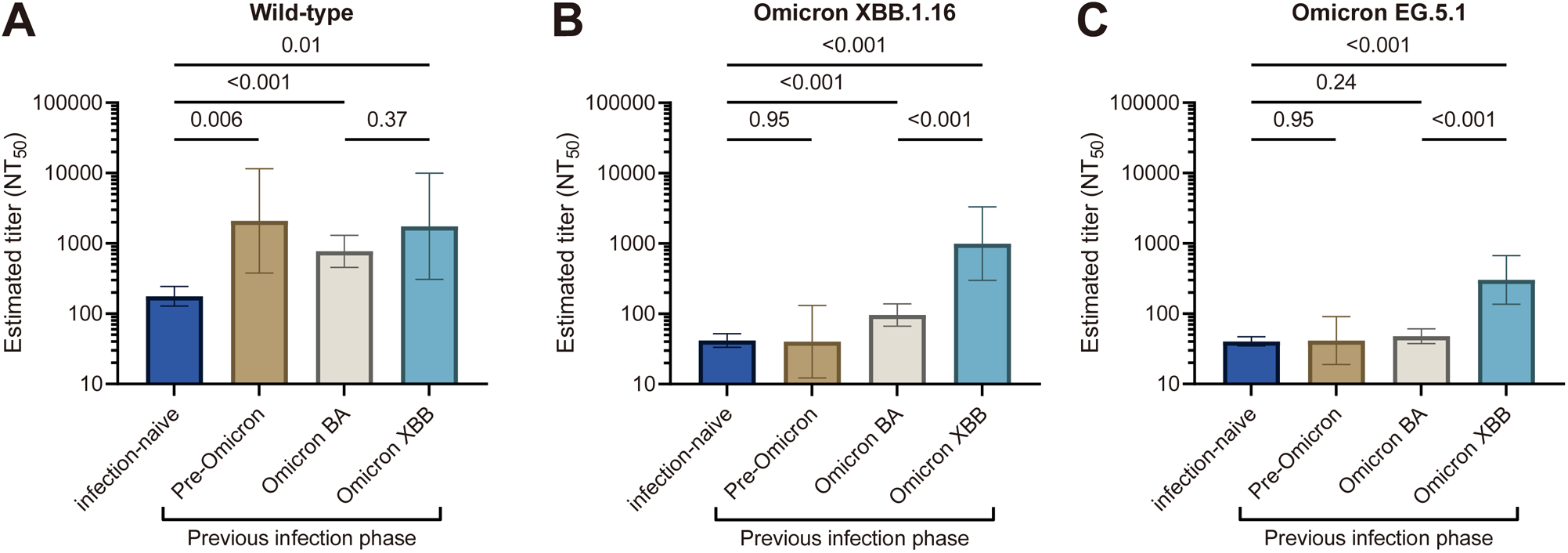
Neutralizing antibody titers across the timing of previous infection. Shown are the geometric mean titers of preinfection-neutralizing antibodies against Wild-type (**A**), Omicron XBB.1.16 (**B**), and EG.5.1 (**C**) across the timing of the previous infection, estimated using a linear regression model while adjusting age, sex, a history of Omicron bivalent vaccination, and the interval between last vaccination and blood sampling. The sample size of infection-naïve, Pre-Omicron, Omicron BA, and Omicron XBB groups are 58, 2, 22, and 2, respectively. Individuals with undiagnosed infection and having neutralizing titers (n=18) were not included in this analysis since their infection timing was unclear. We defined each previous infection phase as follows: Pre-Omicron (February 2020 to December 2021), Omicron BA (January 2022 to March 2023), and Omicron XBB (April 2024 to June 2024). Bars indicate estimated geometric mean titers, and I-shaped bars indicate 95 confidence intervals.

## Discussion

From June to September 2023, when Omicron XBB.1.16 and EG.5.1 were predominantly circulating in Japan, previous infection during the Omicron BA and XBB waves was associated with a 70% and 100% lower risk of subsequent symptomatic SARS-CoV-2 infection, respectively, while the Omicron BA bivalent vaccination was not associated with the risk of infection in a cohort of healthcare workers with three or more doses of vaccination. The preinfection neutralizing antibody titers against Omicron XBB.1.16 and EG.5.1 were lower in infected cases than in the matched controls.

Although the evidence regarding the protection of previous infections and the bivalent vaccine against Omicron XBB.1.16 and EG.5.1 infection is scarce, our findings were similar to a study of U.S. healthcare workers [12]. That study reported that previous Omicron infection was associated with a 60% lower risk of subsequent infection during the dominant waves of Omicron BA.4/5, BQ, or XBB subvariants, whereas the Omicron BA bivalent vaccination was not associated with the risk of infection during the dominant wave of XBB subvariant. Regarding the protection of previous infection, our study has some strengths over the previous study. We classified previous Omicron infection by the subvariant-specific dominant phases (BA or XBB). In addition, we followed subsequent infections restricted to the dominant phase of the XBB.1.16 and EG.5.1. These gaps allowed us to estimate the Omicron BA- and XBB-specific protection against subsequent infections during the XBB.1.16 and EG.5.1 specific-waves, and we found the previous infection during Omicron XBB wave has superior protection against infection during XBB.1.16 and EG.5.1 dominant phase to those during Omicron BA wave (100% vs 70% protection).

In spite of much lower preinfection neutralizing titers against XBB.1.16 and EG.5.1 than those against wild-type, we found that higher neutralizing antibody titers against Omicron XBB.1.16 and EG.5.1 were associated with a lower risk of COVID-19 infection when these variants were dominant. Our findings suggest that variant-specific neutralizing antibody titers could correlate with protection against infection with its variant, even within the low titers range. We also found that the prevalence of those with hybrid immunity (vaccination and previous infection) was lower in cases than in controls (25% vs. 60%), and individuals with hybrid immunity had higher neutralizing titers against XBB.1.16 and EG.5.1 than infection-naïve individuals. Similarly, previous studies reported that neutralizing titers against XBB.1.16 and EG.5.1 were higher in vaccinated individuals with a history of infections than those without [8, 9, 19]. These results confirm that the evidence that hybrid immunity confers better protective humoral immunity than vaccination alone, which has been recognized for the risk of infection with Omicron BA or earlier variants [20, 21], can be extended to the risk of XBB.1.16 and EG.5.1 infection.

This study had several strengths. We rigorously matched cases and controls using a propensity score estimated by several factors potentially associated with SARS-CoV-2 infection risk, including occupational SARS-CoV-2 exposure risk, living arrangements, comorbidities, infection prevention practices, and high infection risk– behaviors. Blood samples for antibody testing were obtained before infection (1 month before the Omicron XBB.1.16 and EG.5 epidemic onset). Previous SARS-CoV-2 infection was determined according to the history of COVID-19 infection and results of anti-N assays, allowing us to identify undiagnosed infections. We measured the neutralizing antibody titers using live viruses. However, limitations also should be acknowledged. We did not conduct active surveillance to detect SARS-CoV-2 infection during the follow-up period. Data on virus strain was not available for the present cases; however, the cases were most likely due to the Omicron XBB variant, which accounted for more than 90% of sequenced COVID-19 samples in Japan during the follow-up (June to September 2023) (**Figure 1**).

## Conclusion

In the era when Omicron XBB.1.16 and EG.5.1 variants were predominant and the Omicron XBB vaccine was still unavailable in Japan, previous Omicron BA or XBB infection, but not Omicron bivalent vaccination, was associated with a lower risk of SARS-CoV-2 symptomatic infection. The preinfection- and live-virus-neutralizing antibody titers against Omicron XBB.1.16 and EG.5.1 were lower in infected cases than in their matched controls. Those with a history of Omicron BA bivalent vaccine had barely detectable neutralizing titers against these variants. Our results highlight the importance of infection prevention practices when the circulating variants had high immune evasion from immunity acquired by existing vaccines.

## Data Availability

All data produced in the present study are available upon reasonable request to the authors.

## Author Contributions

Drs. Yamamoto and Mizoue had full access to all data in the study and took responsibility for the integrity of the data and the accuracy of the data analysis.

Concept and design: Y.S., Matsuda K., Maeda K., M.T., S.H., S.W., O.N.

Acquisition, analysis, or interpretation of data: Y.S., Matsuda K., Maeda K., H.K., O.K., T.T., O.Y., I.N., N.T., T.S.J., K.M., M.T.

Drafting of the manuscript: Y.S., M.T.

Critical revision of the manuscript for important intellectual content: Y.S., Matsuda K., Maeda K., T.S.J., M.T., S.H., A.N., S.W., O.N.

Statistical analysis: Y.S., K.M.

Administrative, technical, or material support: Y.S., Matsuda K, Maeda K, H.K., O.K., T.T., O.Y., I.N., N.T., T.S.J., K.M., M.T., S.H., A.N., S.W., O.N.

Supervision: M.T., O.N.

## Additional Contributions

We thank Mika Shichishima for her contribution to data collection and the staff of the Laboratory Testing Department for their contribution to antibody testing.

