## Supplemental Materials for "Protection of Omicron bivalent vaccine, previous infection, and their induced neutralizing antibodies against symptomatic infection with Omicron XBB.1.16 and EG.5.1"

**Supplemental Text 1.** Propensity score matching algorithm

We performed propensity score matching using a 1:1 matching algorithm without replacement, with a caliper width equal to 0.2 of the standard deviation of the logit of the propensity score, to identify controls with baseline characteristics similar to those of the patients. We estimated the propensity score using multivariable logistic regression, with the incidence of symptomatic SARS-CoV-2 infection as the dependent variable and the following variables as covariates: sex, age, job, occupational risk of SARS-CoV-2 infection, body mass index, coexisting diseases (cancer, cardiovascular diseases, diabetes, hypertension, immunosuppressive diseases, chronic kidney disease, and lung diseases), several infection prevention/risk behaviors, use of tobacco products, frequency of alcohol consumption, number of household members, and children-related living arrangements. To assess the balances between pre-match and post-match, absolute standardized differences were estimated for all baseline covariates before and after matching. Standardized differences <0.1 for a given covariate indicated a relatively small imbalance.

**Supplemental Text 2.** Measurement methods for neutralizing antibodies

The neutralizing activity in the sera of cases and controls was determined by quantifying the serum-mediated suppression of the cytopathic effect (CPE) of each SARS-CoV-2 strain in $HeLa_{hACE2\text{-}TMPRSS2}$ cells obtained from the Japanese Collection of Research Bioresources (JCRB) Cell Bank (Osaka, Japan) [16, 17]. Each serum sample was serially diluted five-fold in a culture medium. The diluted sera were incubated with 100 50% tissue culture infectious dose ($TCID_{50}$) of the virus at 37°C for 20 min (final serum dilution range of 1:40 to 1:25,000). Next, the serum–virus mixtures were inoculated with $HeLa_{hACE2\text{-}TMPRSS2}$ cells ($1.0\times10^4$/well) in 96-well plates. The SARS-CoV-2 strains used in these assays were as follows: a Wuhan wild-type strain ($SARS\text{-}CoV\text{-}2^{05\text{-}2N}$)[23], an Omicron XBB.1.16.5 variant ($SARS\text{-}CoV\text{-}2^{TKYF230030/2023}$, GISAID Accession ID; EPI_ISL_17775017), and Omicron EG.5.1.1 variant ($SARS\text{-}CoV\text{-}2^{TKYnat14564/2023}$, GISAID Accession ID; EPI_ISL_18082364). The levels of CPE observed in SARS-CoV-2–exposed cells were determined using the WST-8 assay with the Cell Counting Kit-8 (Dojindo, Kumamoto, Japan) after culturing the cells for three days. The serum dilution that resulted in 50% inhibition of CPE was defined as the 50% neutralization titer ($NT_{50}$). Each serum sample was tested in duplicate, and the average value was used for analysis. Laboratory technicians were blinded to the case-control status.

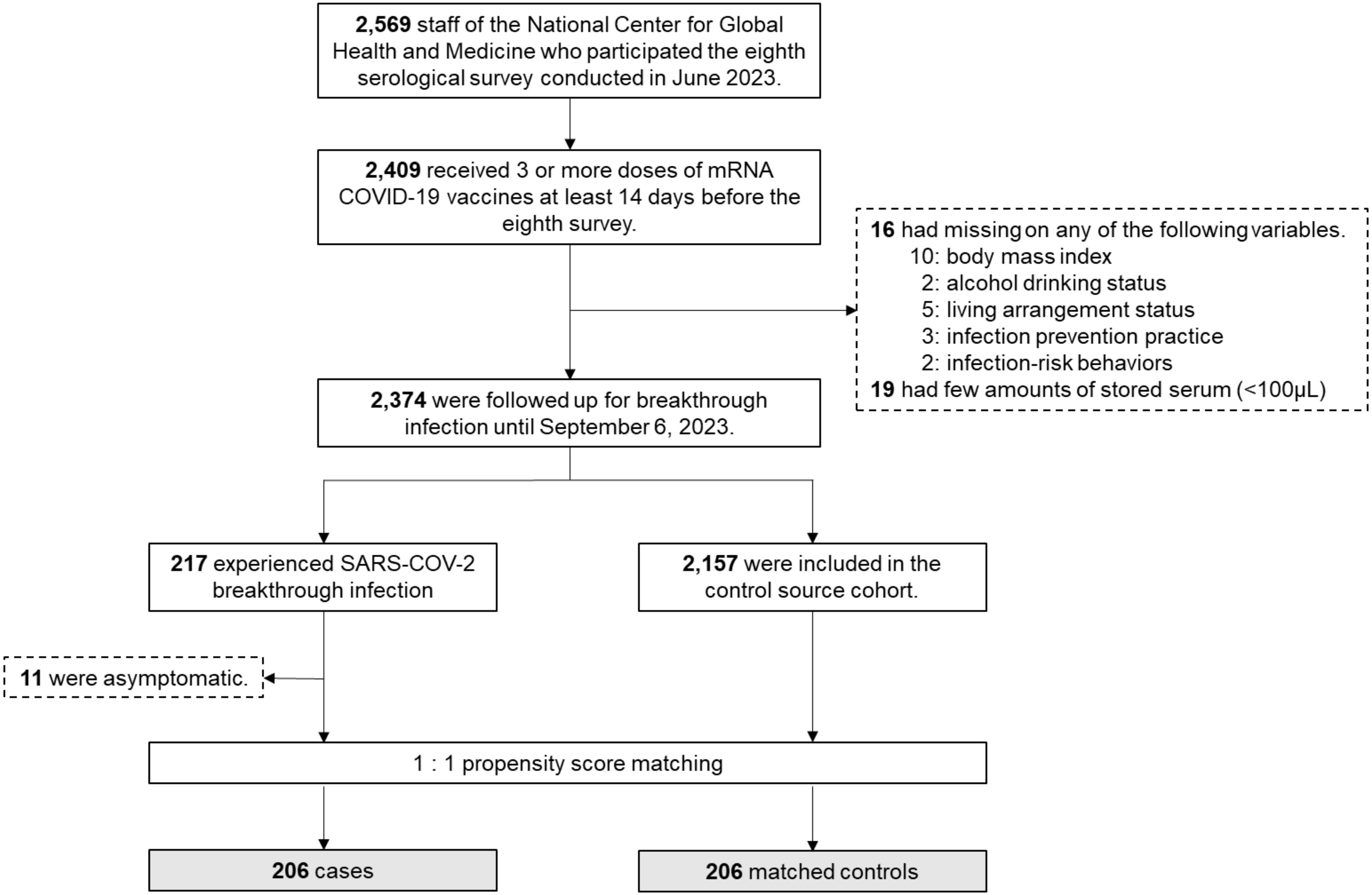

**Supplemental Figure 1.** Case-control selection

Abbreviations: COVID-19, coronavirus disease 2019; SARS-CoV-2, severe acute respiratory syndrome coronavirus 2

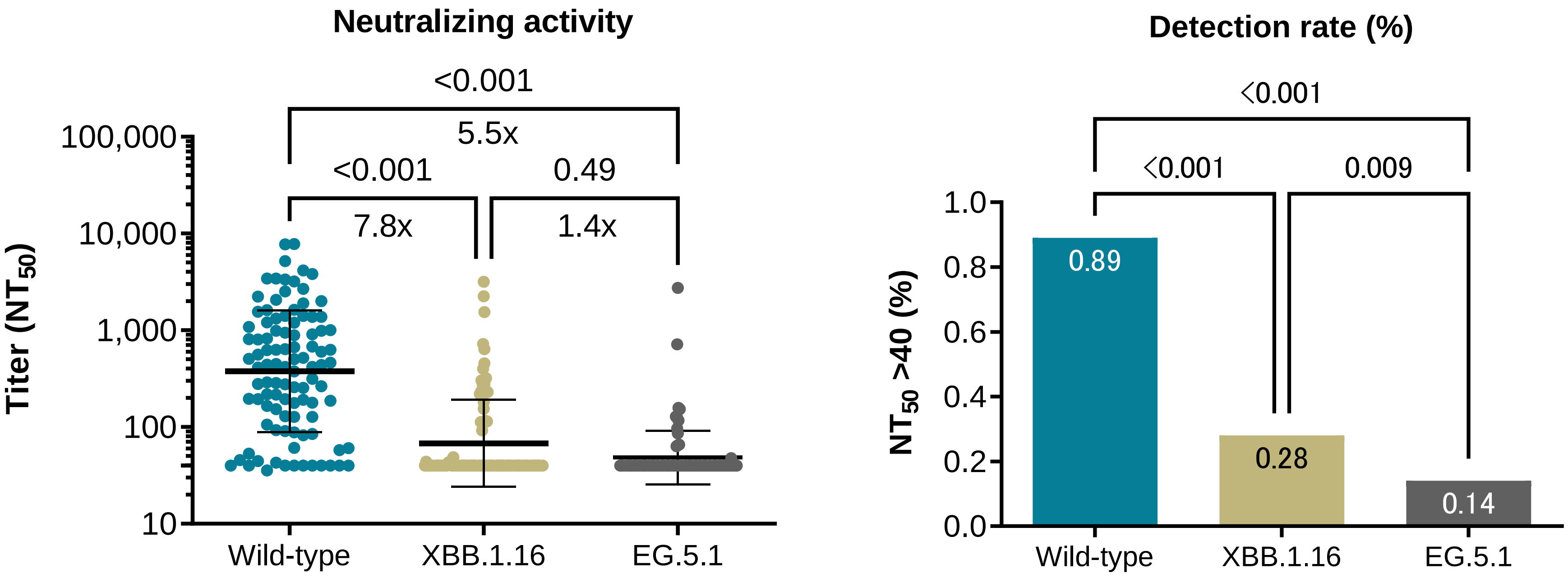

**Supplemental Figure 2.** Cross-reactive neutralization among 100 samples

Comparison of preinfection neutralizing antibody titers against the Wild-type, Omicron XBB.1.16, and Omicron EG.5.1 strains among 100 samples (50 cases and 50 controls). The left panel shows the comparison of neutralizing antibody titers across strains, in which bars indicate geometric mean titers, and I-shaped bars indicate their geometric standard deviations. The right panel shows the comparison of the detection rate of neutralizing antibodies (>40 $NT_{50}$).
